# ANTI - SARS-CoV-2 ANTIBODY SCREENING IN HEALTH CARE WORKERS AND ITS CORRELATION WITH CLINICAL PRESENTATION

**DOI:** 10.1101/2021.07.01.21259884

**Authors:** Suraj Aryal, Sanskriti Pandit, Sushant Pokhrel, Mandira Chhusyabaga, Pabitra Bista, Mahendra Psd. Bhatt, Dharma Datta Subedi, Basista Psd. Rijal

**Author notes:** Corresponding Author: Dr. Basista Psd. Rijal, Professor, Head of Department, Department of Clinical Laboratory Services, Manmohan Memorial Medical College and Teaching Hospital, Kathmandu, Nepal, PO Box: 44600. Co-authors and email:- Suraj Aryal.

## Abstract

**Background:** Antibody titration and the life span of the antibody against SARS-CoV-2 have been found to be determined by the clinical presentation as well. The extent of exposure of health care workers and the general public to SARS-CoV-2 needs to be assessed to monitor the COVID-19 pandemic. Thus, this study is an attempt in assessing the anti-SARs-COV-2 antibody in health care workers.

**Methods:** This laboratory-based cross-sectional study was performed in Manmohan Memorial Medical College and Teaching Hospital, Kathmandu from November 2020 to January 2021. A total of 185 HCWs were screened for anti-SARS-CoV-2 antibody in the blood serum specimens. A structured questionnaire was administered to collect information from HCWs. Anti-SARS-CoV-2 antibody screening was performed using a lateral flow immunoassay. Data were analyzed according to standard statistical methods using SPSS version 20.

**Results:** A total of 185 HCWs were participated in the study, among which 41 (22.2%) of them tested positive for anti-SARS-CoV-2 antibody. Among the 41 individuals who tested positive, 37 of them tested positive for IgG only and 4 of them tested positive for both IgM and IgG antibodies. Presence of history of SARS-CoV-2 infection (p<0.001), presence of flu-like symptoms within the last 6 months (p<0.001), and presence of positive contact history (p=0.002) were statistically significant with antibody screening among HCWs.

**Conclusion:** The burden of SARS-CoV-2 infection among healthcare workers seems to be high and HCWs are at risk of acquiring infection in the workplace. Anti-SARS-CoV-2 antibody screening among health care workers is highly recommended in multiple healthcare settings that can help in monitoring transmission dynamics and evaluation of infection control policies.

## Introduction

Severe acute respiratory syndrome corona virus 2 (SARS-CoV-2) is an enveloped non- segmented positive sense RNA β-corona virus which belongs to the Coronaviridae family (1).It was first diagnosed in December 2019 in a patient showing symptoms of atypical pneumonia from Wuhan, China (2). The virus was found to cause a new disease called corona virus disease 2019 (COVID-19) having high infectious potential (3). Since the first case was identified in China, it has spread globally and it has been declared a pandemic by World Health Organization (WHO) in 11^th^ March, 2020. As of 24^th^ June, 2021, there have been 179,241,734 confirmed cases of Covid-19 globally resulting in 3,889,723 deaths, similarly on the same date the total no. of confirmed cases in Nepal has reached to 629,431 with total no. of 8,918 deaths (4).

Most of the cases of Covid-19 are asymptomatic or with mild symptoms. Individuals characterized as high risk groups (old aged, diabetic, hypertensive, and cancer patients, individuals with pulmonary diseases or disorders) have been found to develop severe clinical presentations leading to multiple organ failure and death (5). The principle of immune response and development of antibodies against SARS-CoV-2 is being studied globally. Infection with SARS-CoV-2 will evoke the development of IgM and IgG antibodies, which is useful in the assessment of immune response following infection. During SARS-CoV-2 infection, IgM and IgG antibodies can be detected in serum within 1-3 weeks after the onset of illness (6). Antibody titration and the life span of antibodies against SARS-CoV-2 have been found to be determined by the clinical presentation as well; short-lived and low antibody titer near the detection limit were found in the cases of mild infections (7).

Healthcare Workers (HCWs) are considered as a highly risk group for SARS-CoV-2 infection. They may acquire infection either from health care setting or from the community. Several studies on the prevalence of SARS-CoV-2 antibodies among HCWs have reported the seroprevalence ranging from 1.6-45.3% (8).According to the study performed by Dimitrios et.al.in a tertiary care centers in North West England, 6% of the health care workers tested positive for SARS-COV-2 IgG antibody (9). The study performed by Deborah et.al.in Belgium showed that 6.4% of healthcare workers tested positive for SARS-COV-2 IgG antibody (10). Studys performed by Mascola et.al.in New York showed that 13.7% of healthcare workers were seropositive for anti-SARS-CoV-2 antibody (11). The extent of exposure of health care workers and the general public to SARS-CoV-2 needs to be assessed to monitor the COVID-19 pandemic. Thus, this study is an attempt in assessing the anti-SARs-COV-2 antibody in health care workers. Exposure to a large number of patients (either symptomatic or asymptomatic) in the hospital for a longer periods of time may be the most common cause of infection for healthcare workers (8, 12). Serological methods for the detection of anti-SARS-CoV-2 antibody could be a useful method for detecting symptomatic and asymptomatic infections (8). Several serological assays for the detection of anti-SARS-CoV-2 antibodies have been in use recently. Serological tests detect the body’s immune response to the infection rather than detecting the virus itself. Thus, this study aims to assess the seroprevalence of anti-SARS-CoV-2 antibody among healthcare workers and determine the association between clinical presentation and antibody screening.

## Methods

This laboratory-based cross-sectional study was performed in Manmohan Memorial Medical College and Teaching Hospital (MMTH), Kathmandu, Nepal, during the period of 3 months (November 2020 to January 2021).

### Inclusion and Exclusion Criteria

Healthcare workers working at MMTH were conveniently selected for the study and HCWs who refused to give informed written consent (n=16) were excluded from the study. All HCWs providing written consent were included in the study.

### Experimental Protocol

Blood sample was collected from HCWs after taking informed written consent by venipuncture following the standard operating protocol. Samples were collected in a serum separator tube (HEBEI XINLE SCI & TECH CO. LTD, China) containing the gel after obtaining the informed and written consent.

Serum was separated from the blood sample using centrifugation. Antibody screening was performed from the serum using SARS-CoV-2 IgM/IgG Antibody Assay Kit (Colloidal Gold Method) (Zybio Inc., China) as per the manufacturer’s instructions (5µl serum was added to the sample well using a pipette followed by the addition of 2 drops of buffer), and the results were read within 10-15 minutes and the results after 15 minutes were considered invalid.

The test was validated by using the serum from the patient with the history of PCR confirmed SARS-CoV-2 infection within the last 3 weeks as a positive control and by using the serum of a healthy individuals without any contact history and without any history of SARS-CoV-2 infection as a negative control.

### Statistical Analysis

Data were collected in Microsoft Excel 2013 and analyzed using SPSS version 20.0 (IBM Corp., Armonk, NY, USA).Age was the only continuous variable defined using median and Inter- quartile range (IQR). Mann-Whitney U test was used for assessing group difference in age. Categorical variables were expressed as frequency rates and percentages. The Chi-square test or Fisher exact test was used as applicable to test for the association between group differences between categorical variables. p-value <0.05 was considered as statistically significant.

## Results

A total of 185 HCWs were participated in the study, among which 41 (22.2%) of them tested positive for anti-SARS-CoV-2 antibody. Among the 41 individuals who tested positive for anti-SARS-CoV-2, 37 were IgG antibody positive while 4 were tested positive for both IgM and IgG antibodies (Figure 1).All 4 individuals who tested positive for both IgM and IgG had the recent history of SARS-CoV-2 infection within a month.

**Figure 1:**
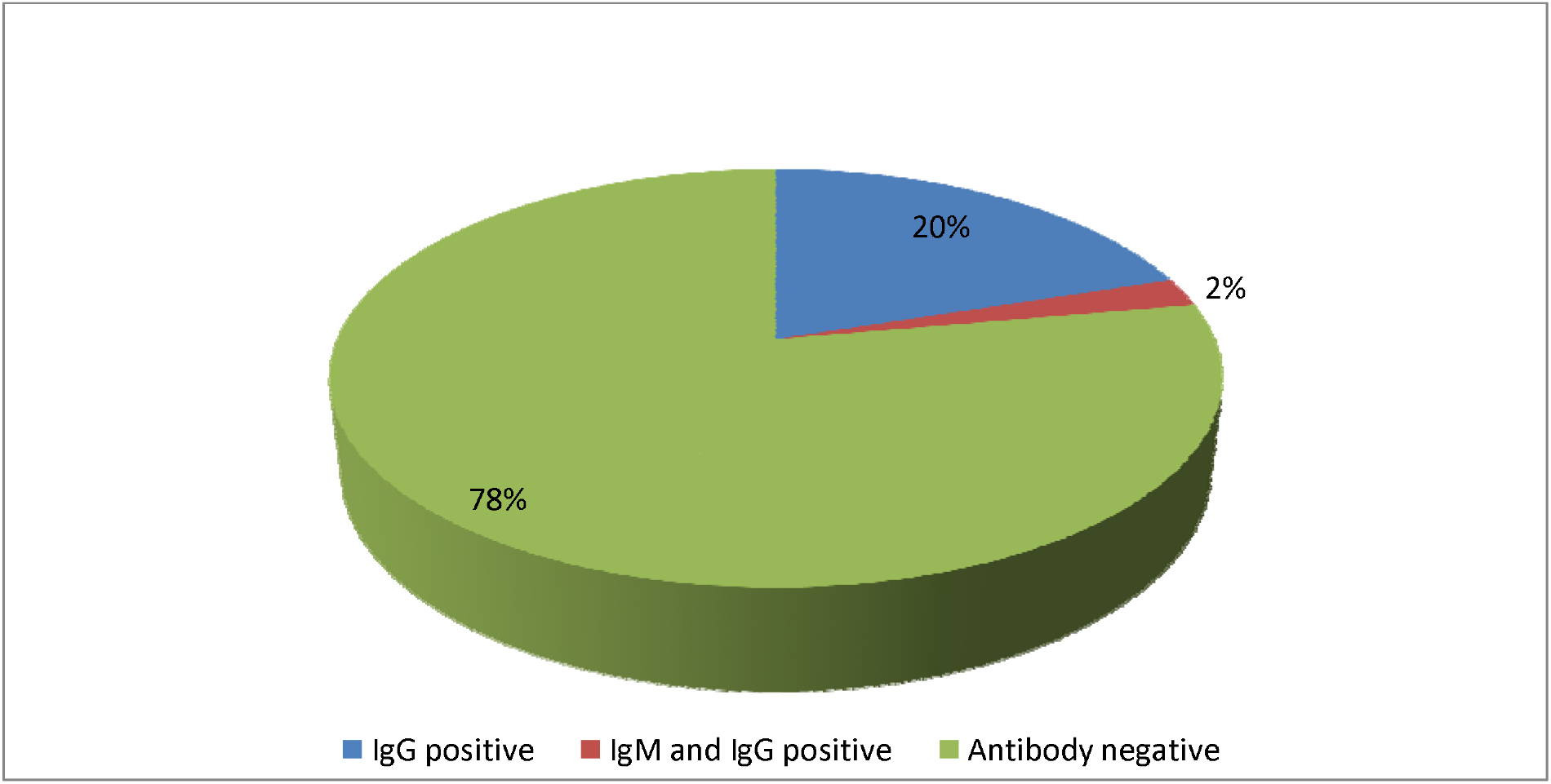
Antibody screening among HCWs.

The median age of HCWs was 27 (IQR 24-36) years and 115 (62.1%) of them were in the age group 20-30. Overall, 52 (28.1%) male and 133 (71.9%) female HCWs participated in the study. Among the 41 individuals, 15 (36.6%) males and 26 (63.4%) females tested positive for antibodies.

The maximum number of individuals who tested positive for antibodies were nurses (39.0%), followed by laboratory professionals (19.5%), administrative staffs (17.1%), doctors (14.6%) and supporting staffs (9.8%) respectively (Table 1).

**Table 1:**
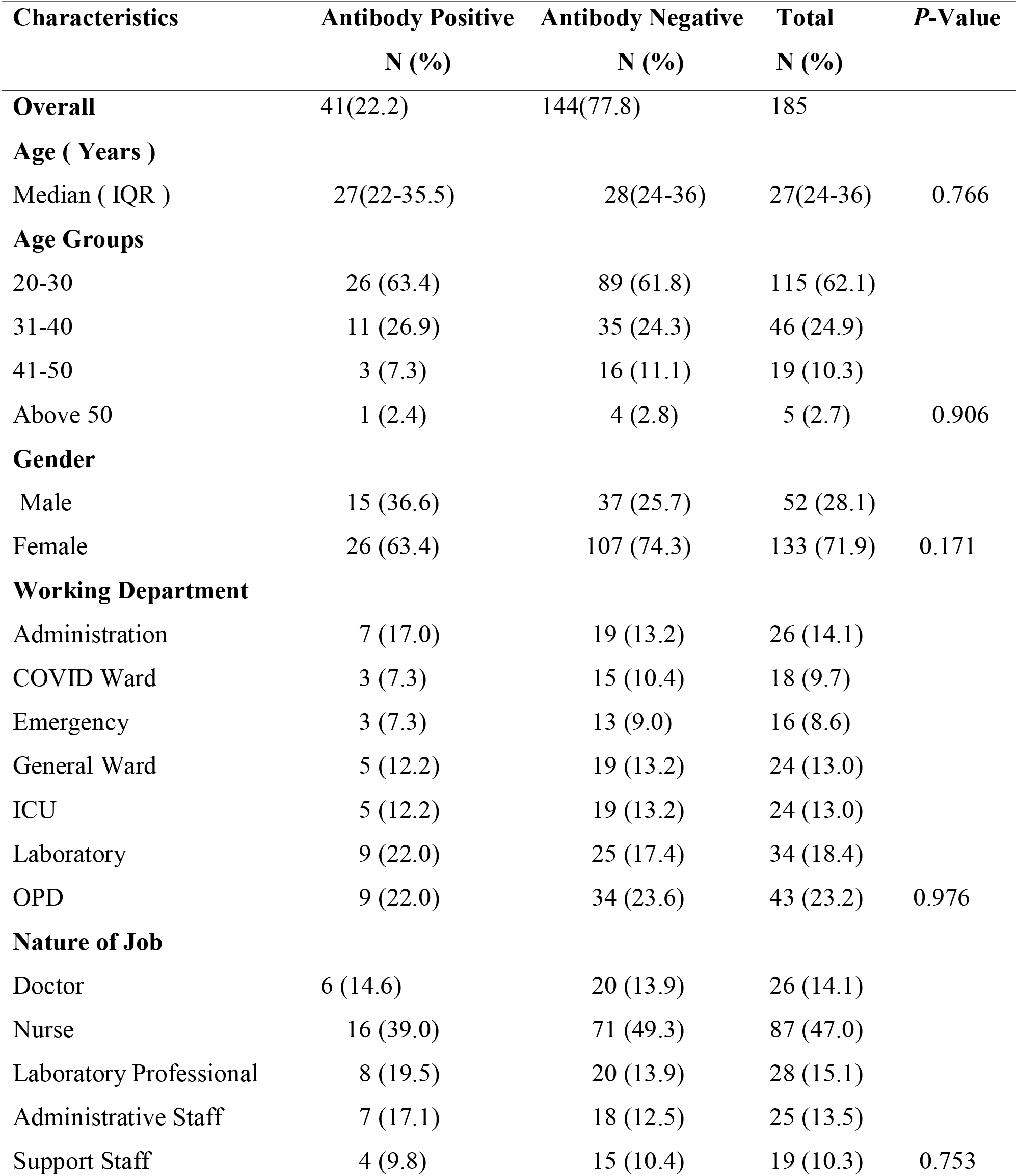
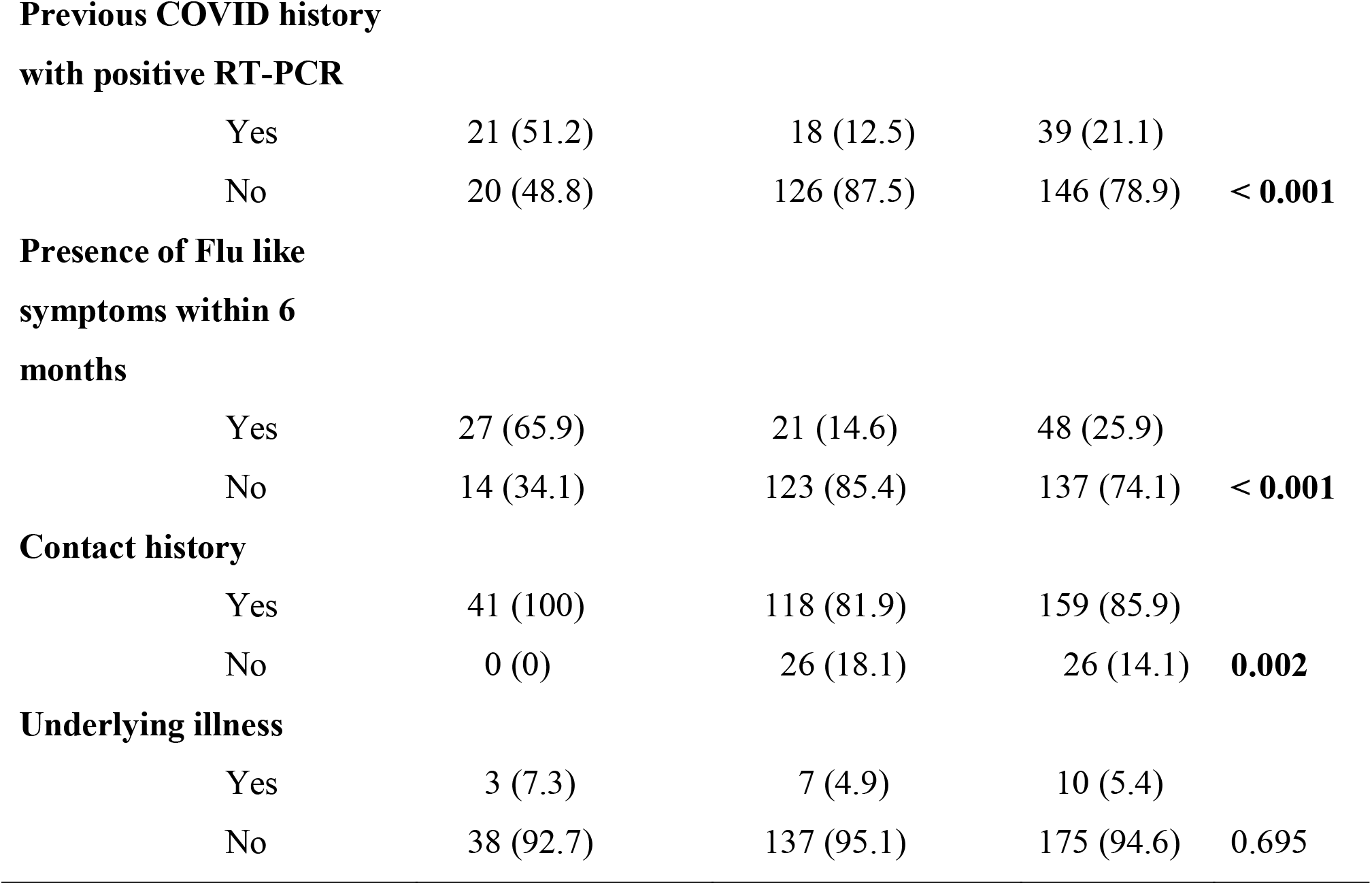
Subject characteristics and group differences.

| **Characteristics** | **Antibody Positive**<br>**N (%)** | **Antibody Negative**<br>**N (%)** | **Total**<br>**N (%)** | **P-Value** |
| --- | --- | --- | --- | --- |
| **Overall** | 41(22.2) | 144(77.8) | 185 |  |
| **Age ( Years )** |  |  |  |  |
| Median ( IQR ) | 27(22-35.5) | 28(24-36) | 27(24-36) | 0.766 |
| **Age Groups** |  |  |  |  |
| 20-30 | 26 (63.4) | 89 (61.8) | 115 (62.1) |  |
| 31-40 | 11 (26.9) | 35 (24.3) | 46 (24.9) |  |
| 41-50 | 3 (7.3) | 16 (11.1) | 19 (10.3) |  |
| Above 50 | 1 (2.4) | 4 (2.8) | 5 (2.7) | 0.906 |
| **Gender** |  |  |  |  |
| Male | 15 (36.6) | 37 (25.7) | 52 (28.1) |  |
| Female | 26 (63.4) | 107 (74.3) | 133 (71.9) | 0.171 |
| **Working Department** |  |  |  |  |
| Administration | 7 (17.0) | 19 (13.2) | 26 (14.1) |  |
| COVID Ward | 3 (7.3) | 15 (10.4) | 18 (9.7) |  |
| Emergency | 3 (7.3) | 13 (9.0) | 16 (8.6) |  |
| General Ward | 5 (12.2) | 19 (13.2) | 24 (13.0) |  |
| ICU | 5 (12.2) | 19 (13.2) | 24 (13.0) |  |
| Laboratory | 9 (22.0) | 25 (17.4) | 34 (18.4) |  |
| OPD | 9 (22.0) | 34 (23.6) | 43 (23.2) | 0.976 |
| **Nature of Job** |  |  |  |  |
| Doctor | 6 (14.6) | 20 (13.9) | 26 (14.1) |  |
| Nurse | 16 (39.0) | 71 (49.3) | 87 (47.0) |  |
| Laboratory Professional | 8 (19.5) | 20 (13.9) | 28 (15.1) |  |
| Administrative Staff | 7 (17.1) | 18 (12.5) | 25 (13.5) |  |
| Support Staff | 4 (9.8) | 15 (10.4) | 19 (10.3) | 0.753 |
| **Previous COVID history with positive RT-PCR** |  |  |  |  |
| Yes | 21 (51.2) | 18 (12.5) | 39 (21.1) |  |
| No | 20 (48.8) | 126 (87.5) | 146 (78.9) | < **0.001** |
| **Presence of Flu like symptoms within 6 months** |  |  |  |  |
| Yes | 27 (65.9) | 21 (14.6) | 48 (25.9) |  |
| No | 14 (34.1) | 123 (85.4) | 137 (74.1) | < **0.001** |
| **Contact history** |  |  |  |  |
| Yes | 41 (100) | 118 (81.9) | 159 (85.9) |  |
| No | 0 (0) | 26 (18.1) | 26 (14.1) | **0.002** |
| **Underlying illness** |  |  |  |  |
| Yes | 3 (7.3) | 7 (4.9) | 10 (5.4) |  |
| No | 38 (92.7) | 137 (95.1) | 175 (94.6) | 0.695 |

Among the 185 individuals tested, 39 (21.1%) individuals had the history of PCR confirmed SARS-CoV-2 infection and among those 39 individuals, only 21 of them tested antibody positive whereas 18 of them tested negative for antibody. All negative 18 subjects had asymptomatic infection and none of them had a history of severe illness.

Overall, 48 (25.9%) individuals had a history of having flu-like symptoms within the last 6 months and 27 of them were found to be antibody positive. Among 27 of the symptomatic antibody positive individuals, body ache, headache, fever, dry cough, and fatigue were the predominant symptoms followed by sore throat, runny nose, anosmia, ageusia and dyspnea (Figure 2).

**Figure 2:**
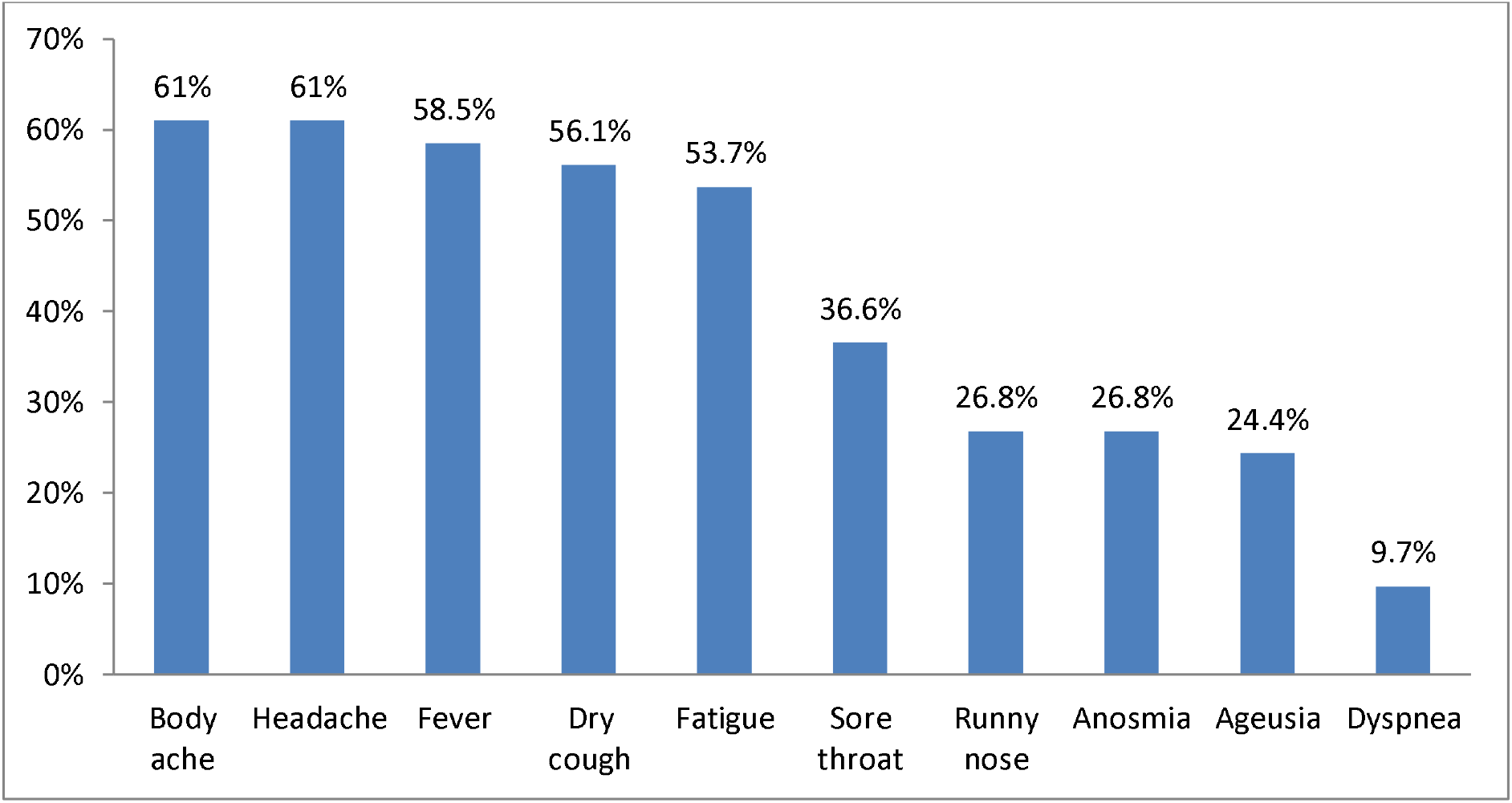
Clinical presentation among HCWs who tested positive for antibodies.

All 41 individuals who tested positive for the antibody had a positive contact history with the SARS-CoV-2 infected patients. Presence of history of SARS-CoV-2 infection (p<0.001), presence of flu-like symptoms within the last 6 months (p<0.001), and presence of positive contact history (p=0.002) were statistically significant with antibody screening among HCWs (Table 1).

## Discussion

COVID-19 is a global pandemic, infecting more than 179 million people around the globe causing death to more than 3.8 million, as of 24^th^ June, 2021. It is believed that almost all immune-competent individuals after being infected with SARS-CoV-2 will develop an immune response against it (6). HCWs are considered as a highly risk group for SARS-CoV-2 infection. They may acquire infection either from health care setting or from the community. Exposure to a large number of patients (either symptomatic or asymptomatic) in the hospital for a longer periods of time may be the most common cause of infection for healthcare workers (8, 9).

Our study revealed significantly higher seroprevalence among healthcare workers compared to previously published reports, ranging prevalence rates from 1.26% to 19.1%. Study performed by Psichogiou Mina et.al. in Greece found the seroprevalence rate to be 1.26 % and mentioned the low burden of COVID-19 in Greece could be the reason for the lower seroprevalence in the study (8). The study performed by Rudberg Ann-Sofie et.al. in Sweden found the seroprevalence rate to be 19.1% (13). Similar study performed by Lombardi Andrea et.al. in Italy revealed the Seroprevalence rate to be 7.6% (14). The higher seroprevalence in our study is corresponding with the 24.4% prevalence rate as reported by Adrian M. et.al. in UK (15). The lack of programs for regular screening of HCWs for SARS-CoV-2 infection, long-term exposure of HCWs in infected cases (symptomatic or asymptomatic), and lack of proper PPE might explain higher seroprevalence in our study (16).

In our study, 39.0% of seropositive HCWs were nurses, suggesting that the seroprevalence is higher among nursing staffs than in other HCWs. This could be due to the reason that nursing staffs are directly involved in patient care and treatment and the participation of nursing staff was higher in this study in comparison to other HCWs. Similarly, the study performed by John T Wilkins et.al.in Chicago had suggested nurses as the highest risk groups for obtaining infection (17) and study performed by Al Maskari et.al. in Oman reported that 38% of the infected HCWs were nurses (12).

Among the study population, 51.2% of seropositive HCWs in our study had the history of RT-PCR confirmed SARS-CoV-2 infection and the remaining 48.8% never obtained a positive RT-PCR results until the time of investigation. Our study showed a significant association between antibody screening and the history of SARS-CoV-2 infection among HCWs, suggesting that HCWs previously infected with SARS-CoV-2 are more likely to be seropositive which is in accordance with the study performed in New York where 93% of PCR positive HCWs were also seropositive (11). Similarly, 40% of seropositive cases in the study performed in Spain did not have the history of SARS-CoV-2 infection and the same study suggested that those infections might have gone undetected in the diagnostic procedure for SARS-CoV-2 infection (16).

65.9% seropositive HCWs had flu-like symptoms within the last 6 months. Although the time duration for which the antibody remained intact following SARS-CoV-2 infection is unclear, the studies performed by L Huillier et.al. in Geneva had shown that the antibody titers donot reduce until 6 months after the infection (18). Studies around the globe like study in Italy (14), study in Spain(16), Study in UK (15) have shown the significant associataion between the positive serology and the presence of symptoms. Body ache and headache were the most common symptoms followed by fever and dry cough among the seropositive HCWs in this study. Studys performed in a large populations of HCWs in United States has reported the presence of at least one symptom among fever, cough, and dyspnea in COVID-19 infection (19). Although body ache, headache, and fever were the commonest symptoms among seropositive, anosmia and ageusia were also reported in 26.8% and 24.4% HCWs, respectively. Lombardi et.al. in Italy found that anosmia, ageusia, and fever were the most common symptoms among SARS-CoV-2 infected HCWs (20). Additionally Rudberg et.al. in Sweden and Gracia Basteiro et.al. in Spain have discussed ageusia and anosmia as the most predictive symptoms of COVID-19 infection in their studies (13, 16). Our study showed lower percentage of seropositive HCWs showing anosmia and ageusia which could be described as variation due to subjects characteristics and geographical variations(14). We found that 34.1% of seropositive HCWs were asymptomatic until the time of investigation since the last 6 months. Similarly, Gracia Basteiro et.al. reported 23.1% seropositive HCWs to be asymptomatic (16). Asymptomatic infection among HCWs made it difficult to identify the infected ones that could degrade the strategies to control the infection, therefore programs for regular screening of HCWs for COVID-19 are essential in controlling the spread of infection.

We tested 185 HCWs in our study, among which 41 were seropositive and all of them had the positive contact history with the COVID infected patient. The positive contact history was significantly associated with the positive serology that suggests that HCWs with positive contact history are more likely to be seropositive, which was in accordance with the findings of Rudberg et.al. where the HCWs with contact to COVID-19 patients had higher seroprevalence than HCWs with contact to non-COVID-19 patient (13). Studys performed by Andrea Lombardi et.al. in Italy also found that HCWs with a positive history of being in contact with COVID-19 infected patients are more likely to be seropositive (14).

This Study had some limitations.This study was time-framed, single center study with a relative small size population. It could have been better if we could use ELISA or CLIA for antibody testing.

## Conclusion

The seroprevalence among healthcare workers was found to be 22.2%. The burden of SARS-CoV-2 infection among healthcare workers seems to be high and HCWs are at risk of acquiring infection in the workplace. High prevalence among HCWs was explained by the long-time exposure of HCWs to SARS-CoV-2 infected patients. The study revealed symptomatic infections are more likely to develop strong antibodies.

The higher seroprevalence in our study could be due to the lack of programs for regular screening of HCWs for SARS-CoV-2 infection, long-term exposure of HCWs in infected cases (symptomatic or asymptomatic), lack of proper PPE. Importantly, during this pandemic, adequate PCR testing and PPE usage was hampered throughout the nation because of the limitation in the availability of testing kits and PPE supply that could have accounted for the higher seroprevalence among HCWs. This state of the problem could highlight the steps to be taken forward to be prepared for such outbreaks, ensuring that the diagnostic capability and PPE supply is adequate.

## Data Availability

All data generated during this study are presented in this paper. The primary raw data will be made available to interested researchers by the first author if requested

## Abbreviations

SARS-CoV-2: Sub Acute Respiratory Syndrome Corona Virus 2
COVID-19: Coronavirus Disease 2019
WHO: World Health Organization
HCWs: Health Care Workers
MMTH: Manmohan Memorial Medical College and Teaching Hospital
SPSS: Statistical Package for Social Sciences
IQR: Inter Quartile Range
ELISA: Enzyme Linked Immuno-sorbent Assay
CLIA: Chemiluminescence Immunoassay
PPE: Personal Protective Equipment

## Acknowledgement

We thank all healthcare workers participating in this study. Our special thanks go to the laboratory staff, management, and officials of Manmohan Memorial Medical College and Teaching Hospital, Kathmandu, for providing the opportunity to carry out this research work.

## Authors’ contribution

BPR, DDS, and MPB - conceived the design of the study, reviewed the literature, performed the necessary interventions including laboratory investigations. SA, SP_1_, MC, and PB- participated in data collection and laboratory procedures. SA and SP_2_ - analyzed data. SA, SP2, and BPR- prepared manuscript. All authors contributed to drafting and critically revising the paper and agree to be accountable for all aspects of the work.

## Ethical Clearance

This research work was approved by the Institutional Review Committee of Manmohan Memorial Institute of Health Sciences (MMIHS-IRC 491), Kathmandu, Nepal. Informed written consent was taken from the healthcare workers before participating inthe study. Data regarding personal information were coded and kept confidential.

## Availability of Data and Materials

All data generated during this study are presented in this paper. The primary raw data will be made available to interested researchers by the corresponding author if requested.

## References

1. Organization WH, Organization WH. Naming the coronavirus disease (COVID-19) and the virus that causes it. 2020.

2. Huang C, Wang Y, Li X, Ren L, Zhao J, Hu Y, et al. Clinical features of patients infected with 2019 novel coronavirus in Wuhan, China. The lancet. 2020;395(10223):497–506.

3. Guo Y-R, Cao Q-D, Hong Z-S, Tan Y-Y, Chen S-D, Jin H-J, et al. The origin, transmission and clinical therapies on coronavirus disease 2019 (COVID-19) outbreak–an update on the status. Military Medical Research. 2020;7(1):1–10.

4. Organization WH. WHO coronavirus disease (COVID-19) Dashboard. Data last updated: 2021//6. 2021.

5. Wendel S, Kutner JM, Machado R, Fontão-Wendel R, Bub C, Fachini R, et al. Screening for SARS-CoV-2 antibodies in convalescent plasma in Brazil: Preliminary lessons from a voluntary convalescent donor program. Transfusion. 2020.

6. Control CfD, Prevention. Interim guidelines for COVID-19 antibody testing. Availavle from: https://wwwcdcgov/coronavirus/2019-ncov/lab/resources/antibody-tests-guidelineshtml [Accessed August 2020]. 2020.

7. Kellam P, Barclay W. The dynamics of humoral immune responses following SARS-CoV-2 infection and the potential for reinfection. Journal of General Virology. 2020:jgv001439.

8. Psichogiou M, Karabinis A, Pavlopoulou ID, Basoulis D, Petsios K, Roussos S, et al. Antibodies against SARS-CoV-2 among health care workers in a country with low burden of COVID-19. PLOS ONE. 2020;15(12):e0243025.

9. Poulikakos D, Sinha S, Kalra PA. SARS-CoV-2 antibody screening in healthcare workers in a tertiary centre in North West England. Journal of Clinical Virology. 2020.

10. Steensels D, Oris E, Coninx L, Nuyens D, Delforge M-L, Vermeersch P, et al. Hospital-wide SARS-CoV-2 antibody screening in 3056 staff in a tertiary center in Belgium. Jama. 2020;324(2):195–7.

11. Moscola J, Sembajwe G, Jarrett M, Farber B, Chang T, McGinn T, et al. Prevalence of SARS-CoV-2 Antibodies in Health Care Personnel in the New York City Area. JAMA. 2020;324(9):893–5.

12. Al Maskari Z, Al Blushi A, Khamis F, Al Tai A, Al Salmi I, Al Harthi H, et al. Characteristics of healthcare workers infected with COVID-19: A cross-sectional observational study. International Journal of Infectious Diseases. 2021 2021/01/01/;102:32-6.

13. Rudberg A-S, Havervall S, Månberg A, Falk AJ, Aguilera K, Ng H, et al. SARS-CoV-2 exposure, symptoms and seroprevalence in health care workers. medRxiv. 2020:2020.06.22.20137646.

14. Lombardi A, Mangioni D, Consonni D, Cariani L, Bono P, Cantù AP, et al. Seroprevalence of anti-SARS-CoV-2 IgG among healthcare workers of a large university hospital in Milan, Lombardy, Italy: a cross-sectional study. BMJ Open. 2021;11(2):e047216.

15. Shields AM, Faustini SE, Perez-Toledo M, Jossi S, Aldera E, Allen JD, et al. SARS-CoV-2 seroconversion in health care workers. medRxiv. 2020:2020.05.18.20105197.

16. Garcia-Basteiro AL, Moncunill G, Tortajada M, Vidal M, Guinovart C, Jiménez A, et al. Seroprevalence of antibodies against SARS-CoV-2 among health care workers in a large Spanish reference hospital. medRxiv. 2020:2020.04.27.20082289.

17. Wilkins JT, Gray EL, Wallia A, Hirschhorn LR, Zembower TR, Ho J, et al. Seroprevalence and Correlates of SARS-CoV-2 Antibodies in Health Care Workers in Chicago. Open Forum Infectious Diseases. 2021;8(1).

18. L’Huillier AG, Meyer B, Andrey DO, Arm-Vernez I, Baggio S, Didierlaurent A, et al. Antibody persistence in the first 6 months following SARS-CoV-2 infection among hospital workers: a prospective longitudinal study. Clinical Microbiology and Infection. 2021.

19. Characteristics of Health Care Personnel with COVID-19 - United States, February 12-April 9, 2020. MMWR Morbidity and mortality weekly report. 2020 Apr 17;69(15):477-81. PubMed PMID: 32298247. Pubmed Central PMCID: PMC7755055 Journal Editors form for disclosure of potential conflicts of interest. No potential conflicts of interest were disclosed. Epub 2020/04/17. eng.

20. Lombardi A, Consonni D, Carugno M, Bozzi G, Mangioni D, Muscatello A, et al. Characteristics of 1573 healthcare workers who underwent nasopharyngeal swab testing for SARS-CoV-2 in Milan, Lombardy, Italy. Clinical Microbiology and Infection. 2020 2020/10/01/;26(10):1413.e9-.e13.

